# Quantifying COVID-19 vaccination hesitancy during early vaccination rollout in Canada

**DOI:** 10.1101/2021.04.29.21256333

**Authors:** Xuyang Tang, Hellen Gelband, Nico Nagelkerke, Isaac I. Bogoch, Patrick Brown, Ed Morawski, Theresa Tam, Angus Reid, Prabhat Jha, for the Action to beat coronavirus/Action pour battre le coronavirus (Ab-C) Study Investigators

## Abstract

**Background:** Understanding vaccination hesitancy during early vaccination rollout in Canada can help the government’s vaccination efforts in education and outreach, which may help eventually achieving herd immunity. This study uses an online survey to assess vaccination hesitancy in population subgroups in Canada.

**Method:** Panel members from the nationally representative Angus Reid Forum were randomly invited to complete an online survey on their experiencing with COVID-19 symptoms and testing, as well as intention to vaccination against COVID-19. Respondents were asked “when a vaccine against the coronavirus becomes available to you, will you get vaccinated or not?” Vaccination hesitancy was defined as choosing “No – I will not get a coronavirus vaccination” as a response.

**Results:** 14,621 panel members (46% male and 53% female) completed the survey. Although the respondents overrepresent age 60+ and higher levels of education, other demographics, the prevalences of smoking, obesity, diabetes and hypertension were comparable to the Canadian national census and health surveys. COVID-19 vaccination hesitancy is relatively low overall (9%). Being a resident of Alberta (predicted probability = 15%), aged 40-59 (OR = 0.87, 0.78 – 0.97, predicted probability = 12%), identifying as a visible minority (OR = 0.56, 0.37 – 0.84, predicted probability = 15%), having some college level education or lower (predicted probability = 14%), or living in households of at least 5 are related to greater vaccination hesitancy (OR = 0.82, 0.76 – 0.88, predicted probability = 13%).

**Conclusion:** Our study enhances the understanding of COVID-19 vaccination hesitancy and identifies key population groups with higher vaccination hesitancy. As the Canadian COVID-19 vaccination effort continues, policymakers may focus outreach, education, and other efforts on these groups, which also represent groups with higher risks for contracting and dying from COVID-19. Furthermore, Canada would need to vaccinate virtually the entire population to reach herd immunity due to its relatively low infection level, and a high vaccination hesitancy would be a major hurdle to achieving that.

## Introduction

Although the Canadian COVID-19 mortality rate is low compared to that in the United States and some European countries, many parts of Canada experienced a significant third wave, driven by more infectious variants and premature lifting of public health measures.^1–3^

Canada started vaccinating healthcare workers and nursing home residents in December 2020, and has prioritized the rest of the population by mortality risk (e.g. age, medical comorbidity) and some provinces further prioritize based on risk of disease acquisition (e.g., essential workers; living in highly impacted regions).^4,5^ Progress in recent weeks has been strong with about 1% of Canadians being vaccinated with at least one dose per day in recent weeks.^6^ Vaccination hesitancy may thwart the effort to reach high vaccine coverage. As a part of the ongoing Action to Beat Coronavirus (Ab-C) study^7^, we report on levels and predictors of self-reported vaccination hesitancy in a reasonably representative sample of Canadians.

## Methods

### Study design and survey

The original study design and participants’ description can be found in earlier reports that examined participants’ symptoms related to COVID-19 and the seroprevalence of SARS-CoV-2 antibodies.^8–10^ Most demographic characteristics of the respondents in the Ab-C study were similar to those from the 2016 Census.^9,11,12^ The older age distribution of the respondents Ab-C versus the Census is a result of intentionally oversampling individuals age 60 and older. Also, the Ab-C cohort has a somewhat higher education level than the national profile (23% with a Bachelor’s degree or higher in Canada and 43% in our sample), hence all analyses are adjusted for education levels.^13^ Nevertheless, the overall demographic distribution of the surveyed panel was similar to the Canadian Census distribution, and the prevalences of smoking, diabetes, hypertension was also similar to national surveys.^14–17^ To learn about vaccine hesitancy, we re-surveyed the original cohort^10^ about their experience with symptoms related to COVID-19 since the last survey, their experience with COVID-19 testing, and their intention to be vaccinated.

### Survey question

Participants were asked “when a vaccine against the coronavirus becomes available to you, will you get vaccinated or not?” The five response choices were: 1. “No – I will not get a coronavirus vaccination,” 2. “Not sure,” 3. “Yes – I will eventually get a vaccination, but I will wait awhile first,” 4. “Yes – I will get a vaccination as soon as one becomes available to me,” 5. “I have already been vaccinated.”

Because the action and opportunity to obtain a vaccine is largely determined by the government’s policy on vaccination eligibility rather than respondents’ own decision making, we assume that those who responded “Yes – I will get a vaccination as soon as one becomes available to me” and those who chose “I have already been vaccinated” have similar levels of intention to vaccinate. Hence, we combined these two groups into one category and created a 4-category variable: 1 = “No – I will not get a coronavirus vaccination,” 2 = “Not sure,” 3 = “Yes – I will eventually get a vaccination, but I will wait awhile first,” 4 = “Yes – I will get a vaccination as soon as one becomes available to me or I have already been vaccinated.”

### Analytical method

Based on previous Ab-C reports, we know that the survey sample is largely representative of the Canadian non-nursing home Census population, but with a smaller proportion with some college education or below and a higher proportion with a bachelor degree or higher. To adjust for this, we applied the same survey weights as in prior reports in the ordered logit models that examine the odds of having stronger intention to vaccinate.

Prior to calculating odds ratios, parallel line assumptions (also known as the proportional odds assumption, i.e. the relationship between demographics and different thresholds of the outcome is proportional) were tested for each demographic factor at the 0.05 level within the survey setting, and all variables did meet the parallel line assumption. The overall model also meets the parallel line assumption with an adjusted Wald test. Sensitivity analysis that a used weighted generalized ordered logit testing the parallel line assumption and relaxing the assumption if variables fail to meet it had shown that the results were the same as the weighted ordered logit where the parallel line assumption held (data not shown). Marginal effects of the weighted ordered logit were also calculated to show the predicted probabilities of each vaccination intention level by demographic group. All analyses were done in Stata 16.^18^

### Ethics

The Ab-C study was approved by the IRB of St. Michael’s Hospital, Unity Health Toronto.

## Results

Between January and March 2021, 14,621 participants out of 20,359 polled responded to the survey. The respondents are demographically comparable to the Canadian population in terms of age, gender, and household size. (Table 1), with the exception of the proportion of the Indigenous group (8.9% in the Ab-C and 5% in Canada) and as mentioned above, educational attainment (43% bachelor’s degree or higher in Ab-C and 23% in Canada). In contrast, Ab-C participants also have similar prevalences of obesity, diabetes and hypertension as the national population.

**Table 1.** Unweighted survey sample characteristics (n = 14,621)

|  | Canada | Ab-C | Ab-C respondents who "Will not vaccinate" by demographics |  |  |
| --- | --- | --- | --- | --- | --- |
|  | % | n | % | n | row % |
| **Total** |  | 14,621 | 100% | 1355 | 9.3% |
| **Province** |  |  |  |  |  |
| Ontario | 38% | 5,766 | 39.4% | 448 | 7.8% |
| British Columbia | 13% | 2,761 | 18.9% | 199 | 7.2% |
| Quebec | 23% | 2,299 | 15.7% | 191 | 8.3% |
| Alberta | 12% | 1,839 | 12.6% | 301 | 16.4% |
| Manitoba & Saskatchewan | 7% | 1,027 | 7% | 142 | 13.8% |
| Atlantic provinces | 7% | 929 | 6.4% | 74 | 8% |
| **Age groups** |  |  |  |  |  |
| 18 to 39 years | 49% | 4,270 | 29.2% | 396 | 9.3% |
| 40 to 59 years | 28% | 5,106 | 34.9% | 590 | 11.6% |
| 60 to 69 years | 12% | 3,472 | 23.7% | 257 | 7.4% |
| 70 years or older | 11% | 1,773 | 12.1% | 112 | 6.3% |
| **Sex** |  |  |  |  |  |
| Male | 49% | 6,742 | 46.1% | 736 | 10.9% |
| Female | 51% | 7,777 | 53.2% | 602 | 7.7% |
| Prefer to self-describe |  | 102 | 0.7% | 17 | 16.7% |
| **Education** |  |  |  |  |  |
| Some college or lower | 45% | 3,402 | 23.3% | 468 | 13.8% |
| Some university | 32% | 4,935 | 33.8% | 564 | 11.4% |
| Bachelor or higher | 23% | 6,284 | 43% | 323 | 5.1% |
| **Visible minority** |  |  |  |  |  |
| No |  | 12,526 | 85.7% | 1,098 | 8.8% |
| Yes | 22% | 2,095 | 14.3% | 257 | 12.3% |
| **Ethnicity** |  |  |  |  |  |
| Not Indigenous |  | 13,317 | 91.1% | 1,164 | 8.7% |
| Indigenous | 5% | 1,304 | 8.9% | 191 | 14.6% |
| **Household size** |  |  |  |  |  |
| One person | 28% | 2,678 | 18.3% | 262 | 9.8% |
| Two people | 34% | 6,315 | 43.2% | 501 | 7.9% |
| Three people | 15% | 2,415 | 16.5% | 219 | 9.1% |
| Four people | 14% | 2,163 | 14.8% | 204 | 9.4% |
| Five or more people | 8% | 1,050 | 7.2% | 169 | 16.1% |
| **Weight status** |  |  |  |  |  |
| Under or normal weight | 37% | 4,318 | 29.5% | 396 | 9.2% |
| Overweight | 37% | 4,716 | 32.3% | 447 | 9.5% |
| Obese | 27% | 3,914 | 26.8% | 337 | 8.6% |
| Unknown |  | 1,673 | 11.4% | 175 | 10.5% |
| **Smoking status** |  |  |  |  |  |
| Never | 46% | 7,310 | 50% | 632 | 8.6% |
| Former or current | 54% | 7,063 | 48.3% | 693 | 9.8% |
| Unknown |  | 248 | 1.7% | 30 | 12.1% |
| **Diabetic** |  |  |  |  |  |
| No |  | 13,064 | 89.4% | 1,222 | 9.4% |
| Yes | 9% | 1,424 | 9.7% | 116 | 8.1% |
| Unknown |  | 133 | 0.9% | 17 | 12.8% |
| **Hypertensive** |  |  |  |  |  |
| No |  | 10,419 | 71.3% | 1,026 | 9.8% |
| Yes | 23% | 3,980 | 27.2% | 299 | 7.5% |
| Unknown |  | 222 | 1.5% | 30 | 13.5% |
| **Note** |  |  |  |  |  |
| British Columbia includes Yukon. Manitoba & Saskatchewan include Northwest Territories. |  |  |  |  |  |

Sources:
- a. Census Canada 2016<sup>11-13, 19</sup> - b. Health Fact Sheets<sup>14, 15</sup> - c. Health Reports<sup>16</sup> - d. Public Health Agency of Canada<sup>17</sup>

Among Ab-C participants, 9% have no intention to vaccinate. (Table 1) Alberta (16%) and other Prairie provinces (14%) have the highest proportion who do not intend to vaccinate compared to less than 10% in other provinces. Participants aged 40-59 have the highest vaccination hesitancy, with 11.6% reporting no intention to vaccinate, compared to less than 10% in other age groups. Men have higher vaccination hesitancy (11%%) than women (8%), and those with some college level education or less have stronger vaccination hesitancy (14%) than participants with other levels of education. Other groups with stronger than average vaccination hesitancy include those identifying themselves as visible minority (12%), Indigenous (15%), and those living in households of five or more people (16%).

Table 2 presents participants’ weighted odds ratios of vaccination intention and the predicted probabilities to not vaccinate for each demographic group. Compared to the reference groups, the demographics with lower odds of having strong intention to vaccinate (and thus higher vaccination hesitancy) include age 40-59 years (OR = 0.87, 0.78 – 0.97, predicted probability = 12%), being a visible minority (OR = 0.56, 0.37 – 0.84, predicted probability = 15%), and belonging to a household of five or more people (OR = 0.82, 0.76 – 0.88, predicted probability = 13%). Being from Alberta and having some college level education or lower also have high predicted probability of vaccination hesitancy at 15% and 14%, respectively. The predicted probability of unwillingness for key demographic groups are also displayed in Figure 1.

**Table 2.** Results of the weighted ordered logit for intention to vaccinate and predicted probability for vaccination hesitancy

|  | n | Weighted OR | Weighted 95% CI | Prob | SE |
| --- | --- | --- | --- | --- | --- |
| **Province** |  |  |  |  |  |
| Ontario | 5,766 | Ref |  | 0.092** | (0.012) |
| British Columbia | 2,761 | 1.09 | (0.67, 1.77) | 0.085** | (0.017) |
| Quebec | 2,299 | 1.01 | (0.72, 1.43) | 0.091** | (0.014) |
| Alberta | 1,839 | 0.58 | (0.17, 2.01) | 0.146* | (0.047) |
| Manitoba & Saskatchewan | 1,027 | 0.70 | (0.21, 2.28) | 0.126* | (0.031) |
| Atlantic provinces | 929 | 0.95 | (0.52, 1.74) | 0.096** | (0.014) |
| **Age groups** |  |  |  |  |  |
| 18 to 39 years | 4,270 | Ref |  | 0.107** | (0.022) |
| 40 to 59 years | 5,106 | 0.87** | (0.78, 0.97) | 0.120** | (0.022) |
| 60 to 69 years | 3,472 | 1.41** | (1.03, 1.92) | 0.079** | (0.012) |
| 70 years or older | 1,773 | 1.75*** | (1.50, 2.04) | 0.065** | (0.012) |
| **Sex** |  |  |  |  |  |
| Male | 6,742 | Ref |  | 0.110** | (0.020) |
| Female | 7,777 | 1.23* | (0.97, 1.56) | 0.091** | (0.017) |
| Prefer to self-describe | 102 | 0.76 | (0.38, 1.53) | 0.138* | (0.037) |
| **Education** |  |  |  |  |  |
| Some college or lower | 3,402 | Ref |  | 0.140** | (0.027) |
| Some university | 4,935 | 1.33** | (1.10, 1.62) | 0.110** | (0.020) |
| Bachelor or higher | 6,284 | 2.83*** | (1.96, 4.09) | 0.056** | (0.008) |
| **Visible minority** |  |  |  |  |  |
| No | 12,526 | Ref |  | 0.092** | (0.018) |
| Yes | 2,095 | 0.56** | (0.37, 0.84) | 0.150** | (0.016) |
| **Ethnicity** |  |  |  |  |  |
| Not Indigenous | 13,317 | Ref |  | 0.100** | (0.018) |
| Indigenous | 1,304 | 0.95 | (0.86, 1.05) | 0.104** | (0.017) |
| **Household size** |  |  |  |  |  |
| One person | 2,678 | Ref |  | 0.113** | (0.024) |
| Two people | 6,315 | 1.28** | (1.09, 1.50) | 0.091** | (0.017) |
| Three people | 2,415 | 1.19 | (0.87, 1.62) | 0.097** | (0.015) |
| Four people | 2,163 | 1.20 | (0.82, 1.75) | 0.097** | (0.013) |
| Five or more people | 1,050 | 0.82*** | (0.76, 0.88) | 0.134** | (0.028) |
| **Weight status** |  |  |  |  |  |
| Under or normal weight | 4,318 | Ref |  | 0.107** | (0.016) |
| Overweight | 4,716 | 1.11 | (0.75, 1.63) | 0.098** | (0.021) |
| Obese | 3,914 | 1.18** | (1.05, 1.34) | 0.092** | (0.016) |
| Unknown | 1,673 | 0.93 | (0.59, 1.48) | 0.113** | (0.023) |
| **Smoking status** |  |  |  |  |  |
| Never | 7,310 | Ref |  | 0.100** | (0.019) |
| Former or current | 7,063 | 0.99 | (0.85, 1.15) | 0.101** | (0.017) |
| Unknown | 248 | 0.83 | (0.57, 1.22) | 0.117** | (0.025) |
| **Diabetic** |  |  |  |  |  |
| No | 13,064 | Ref |  | 0.101** | (0.018) |
| Yes | 1,424 | 1.11 | (0.85, 1.44) | 0.092** | (0.021) |
| Unknown | 133 | 0.77 | (0.32, 1.85) | 0.126* | (0.032) |
| **Hypertensive** |  |  |  |  |  |
| No | 10,419 | Ref |  | 0.104** | (0.018) |
| Yes | 3,980 | 1.21** | (1.09, 1.33) | 0.088** | (0.018) |
| Unknown | 222 | 0.72* | (0.47, 1.10) | 0.137** | (0.016) |
| Notes: |  |  |  |  |  |
| British Columbia includes Yukon. Manitoba & Saskatchewan include Northwest Territories. |  |  |  |  |  |
| *** p<0.01, ** p<0.05, * p<0.1 |  |  |  |  |  |

Notes:

**Figure 1.**
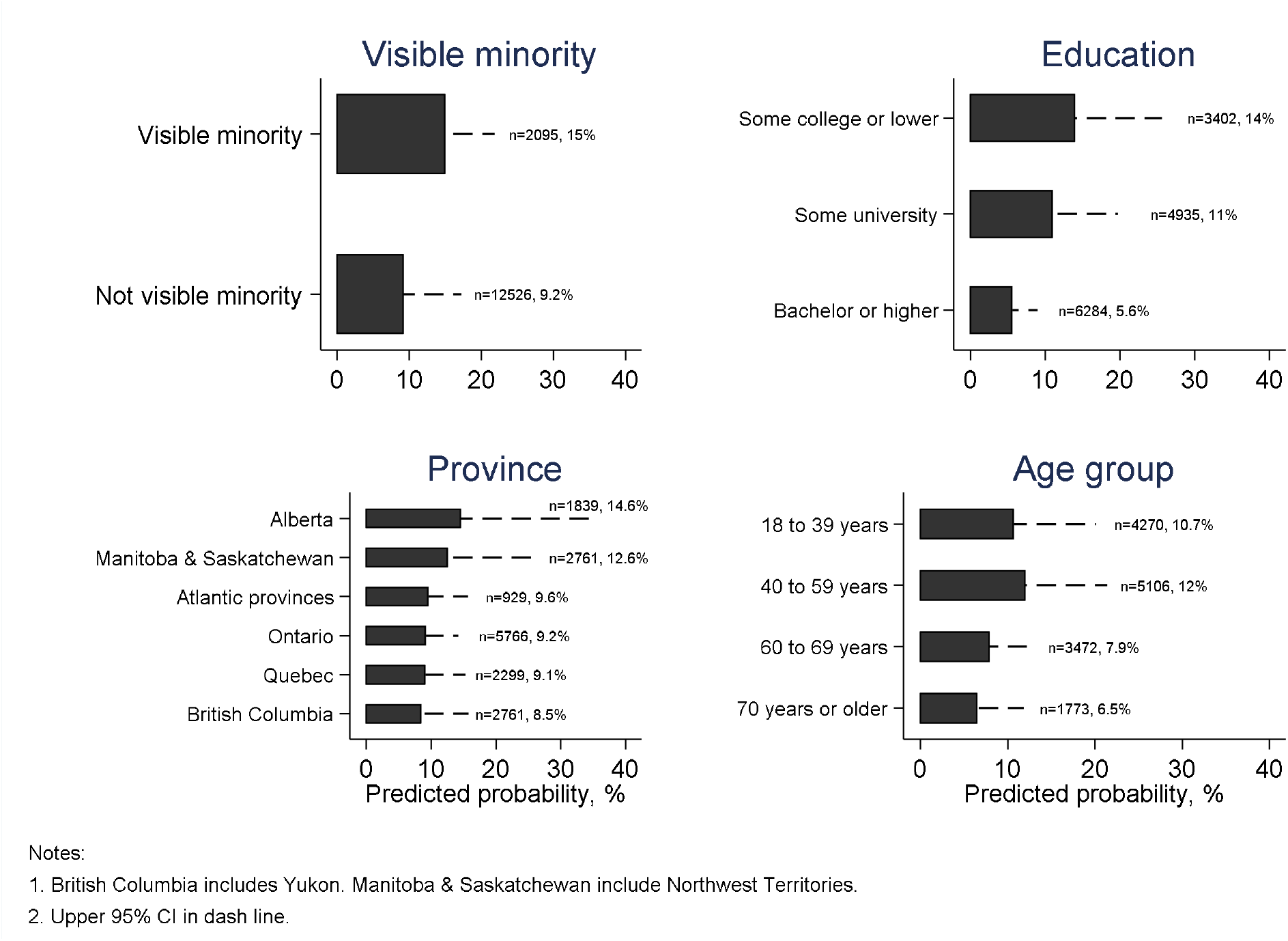
Probabilities of unwillingness to vaccinate for key demographic groups among participants in the Ab-C study

Furthermore, anyone who identifies with 2 of the 3 demographics with the highest predicted probability for vaccination hesitancy (i.e. being from Alberta, having some college education or lower, being a visible minority) has a predicted probability of 2% for having no intention to vaccinate (0.21 and weighted n = 238 for from Alberta and being a visible minority, 2% and weighted n = 631 for being a visible minority and having some college level education or lower, 2% and weighted n = 600 for from Alberta and having some college level education or lower). For anyone who is from Alberta with some college level education or lower and identifies as a visible minority, the predicted probability of having no intention to vaccinate is 3%, although the sample size is very small (weighted n = 89).

## Discussion and next steps

More than 9% of 14,621 participants from the Ab-C study reported having no intention to vaccinate against COVID-19. Being a resident of Alberta, aged 40-59, identifying as a visible minority, having some college level education or lower, or living in households of at least 5 are related to greater vaccination hesitancy. The key differences between the Canadian population and the Ab-C cohort are the oversampling of Forum panel members at least 60 years of age and the high proportion of respondents with a Bachelor’s degree or higher. Both groups have lower vaccination hesitancy compared to their counter parts in the Ab-C survey, making the overall 9% vaccination hesitancy a conservative estimate.

Applying the 9% vaccination hesitancy to Canada as a whole, the approximately 3 million adult Canadians hesitant to vaccinate may play an important role in continuing viral transmission. This population may interact with the approximate 7.2 million Canadians below age 18,^19^ whom are not likely to be vaccinated for the foreseeable future. Moreover, vaccine effectiveness is about 90% for most of the vaccines available in Canada,^20^ which implies that about 3 million vaccinated people don’t have the full immunity. Collectively, these represent over a third of the total population. Within particular sub-groups, such as visible minorities, greater proportions of vaccine hesitancy paired with exposure to children might play a role in fueling sub-waves of transmission within local network.^21,22^ Thus, reducing vaccine hesitancy overall (and eventually vaccinating children) are important.

As the Canadian COVID-19 vaccination effort continues, policymakers may focus outreach, education, and other efforts on these groups, which also represent groups with higher risks for contracting and dying from COVID-19.^23^ As Canada’s vaccination campaign continues, with a goal of vaccinating the majority of the population by September 2021,^4^ we will periodically re-survey the Ab-C cohort and report on vaccination hesitancy and status.

## Data Availability

Data will be made available upon request pending approval from the Unity Health Toronto
Ethics Review Board and the COVID-19 Immunity Task Force.

